# IDENTIFYING ASSOCIATIONS BETWEEN GENETIC CONDITIONS IN OFFSPRING AND PREGNANCY HEALTH COMPLICATIONS

**DOI:** 10.1101/2023.01.17.23284690

**Authors:** Rebecca A. Gibson, Bijan Abar, Lily Elman, Kyle M. Walsh, Jillian H. Hurst, Jennifer L. Cohen

## Abstract

**Background:** Prior research has identified associations between pregnancy complications and specific fetal genetic diagnoses in the offspring.

**Objective:** The purpose of this study is to provide a large-scale investigation of associations between infant genetic diagnoses and pregnancy complications.

**Methods:** Retrospective chart review of maternal-infant dyads born at Duke University (2013-2021) identified 236 mothers who had infants with genetic diseases (exposed group) and 472 matched dyads without concern for newborn genetic disease (unexposed group). Logistic regression examined associations between gestational factors and infant genetic diseases.

**Results:** The risk of gestational diabetes was elevated among mothers of infants with aneuploidy (RR=1.65; 95% CI 1.05,2.61). The risk of preeclampsia was elevated among mothers of infants with imprinting conditions (RR=5.08; 95% CI 2.41,10.73). The presence of any infant genetic disease was associated with an increase in placental disorders (RR=1.98; 95% CI 1.61,2.43), including a 2.25-fold increase in placental infarction (RR=2.25; 95% CI 1.72,2.93).

**Conclusions:** These findings provide insight into relationships between fetal genetic diseases and gestational complications and may guide identification of pregnancies that would benefit from expanded prenatal and antenatal genetic screening for earlier identification and treatment of genetic conditions.

**SYNPOSIS:** *Study question:* Are there associations between genetic conditions in offspring and pregnancy health complications?

*What’s already known:* Prior research has identified associations between pregnancy complications and specific fetal genetic diseases, including pregnancy-related hypertension/preeclampsia in association with *EP300*-related Rubinstein-Taybi syndrome.

*What this study adds:* This retrospective cohort study provides the first comprehensive investigation into potential links between offspring genetic diagnoses and pregnancy complications, identifying associations between aneuploidy and gestational diabetes, imprinting disorders and preeclampsia, and fetal genetic conditions and specific placental abnormalities. These findings can inform future medical management of pregnancies affected by a fetal genetic condition and help guide the future development of algorithms to prospectively identify parent/infant dyads who may benefit from expanded prenatal and neonatal genetic testing.

## BACKGROUND

The etiologies of complications during pregnancy are complex and remain incompletely understood, but there is accumulating evidence that fetal genetic features may contribute to pregnancy complications. In particular, prior research has identified several associations between pregnancy complications and individual fetal genetic diseases. Infants with numeric sex chromosome defects, including Klinefelter Syndrome and Triple X Syndrome, are more likely to be born to individuals experiencing gestational diabetes.^1,2^ Other studies have identified associations between preeclampsia or gestational hypertension and fetal aneuploidies including Trisomy 13, Trisomy 18, and Trisomy 21 (OMIM # 190685),^3–6^ though these findings have not been consistently replicated.^7,8^ More recently, several monogenic conditions including *EP300*-related Rubenstein Taybi Syndrome (OMIM # 613684), mitochondrial trifunctional protein deficiency (TFP) (OMIM # 609015), ^9,10^ Long-chain 3-hydroxyacyl-CoA dehydrogenase (LCHAD) deficiency (OMIM # 609016),^11–13^ and fatty acid oxidation conditions (FAOD)^14^ have been observed at higher frequencies among children born to individuals with preeclampsia, placental disorders, and gestational liver disease, among other complications during pregnancy. Importantly, these studies have primarily focused on describing pregnancy complications associated with a specific fetal genetic diagnosis.

To date, there have been few broad investigations of potential associations between fetal or infant genetic diagnoses and pregnancy complications. Such an analysis could identify patterns of association between fetal and pregnancy conditions that may help identify mother-infant dyads at risk of poor pregnancy outcomes, help to better elucidate the molecular pathways underlying common pregnancy complications, and guide prospective identification of pregnancies that would benefit from early and/or expanded genetic screening. We therefore conducted a chart review of 708 mother-infant dyads, including 236 dyads with a postnatal infant genetic disease diagnosis (exposed) and 472 matched dyads without genetic disease diagnosis nor a concern for genetic disease (unexposed). Using data derived from this chart review, we evaluated associations between different categories of fetal genetic disease and pregnancy complications.

## METHODS

### Data Collection

We conducted a retrospective study through evaluation of electronic health records (EHR) data at Duke University Health System (DUHS), located in central North Carolina. DUHS is a large, integrated health system consisting of three hospitals and over 100 outpatient clinics. This retrospective study was deemed exempt from review by the DUHS Institutional Review Board (protocol number Pro00106469). All study activities were conducted in accordance with the Declaration of Helsinki.

Electronic health records (EHR) were reviewed and selected data elements were abstracted into a REDCap database^15^ for all mothers and associated infants who were born at DUHS-affiliated hospitals between July 2013 and January 2021. Records were reviewed for identification of inpatient genetic consult or genetic testing in the infant, and for subsequent confirmation of infant genetic diagnosis (a proxy for exposure to a fetal genetic condition). Based on the EHR review, we identified 236 dyads in which the child had a postnatally confirmed genetic diagnosis (exposed group) and 472 matched dyads in which the children did not receive a confirmed genetic diagnosis, were not consulted on by genetics, and did not receive genetic testing (unexposed). Unexposed and exposed dyads were matched at a 2:1 ratio by maternal age at delivery, patient-reported race and ethnicity, parity, and infant sex (Table 1). The unexposed group was filtered for removal of secondary maternal records in the case of a twin gestation and filtered for removal of dyads in which a child did not receive genetic services but was reported to have multiple congenital anomalies. Mothers could be included twice in the dataset if they had two or more distinct pregnancies during the study period.

**Table 1.** Demographics of exposed and unexposed groups. The exposed group includes 236 mother-infant dyads whose child was found to have a confirmed genetic diagnosis. The unexposed group includes 472 mother-infant dyads whose child did not have a suspected or confirmed genetic disease. Unexposed dyads were matched to exposed dyads for maternal age at delivery, race/ethnicity, parity, and infant sex in a 2:1 unexposed to exposed ratio.

|  | Unexposed<br>(n=472) | Exposed<br>(Genetic<br>Disease)<br>(n=236) | Overall<br>(n=708) | p value<br>(<0.05) |
| --- | --- | --- | --- | --- |
| **Ethnicity** |  |  |  | 0.23 |
| Hispanic | 74 (15.7%) | 37 (15.7%) | 111 (15.7%) |  |
| Not Hispanic/Latino | 386 (81.8%) | 193 (81.8%) | 579 (81.8%) |  |
| Not Reported/Declined | 12 (2.5%) | 6 (2.5%) | 18 (2.5%) |  |
| **Race** |  |  |  | 0.90 |
| Asian | 9 (1.9%) | 6 (2.5%) | 15 (2.1%) |  |
| Black | 161 (34.1%) | 81 (34.3%) | 242 (34.2%) |  |
| Not Reported/Declined | 14 (3.0%) | 6 (2.5%) | 20 (2.8%) |  |
| Other | 76 (16.1%) | 39 (16.5%) | 115 (16.2%) |  |
| White | 212 (44.9%) | 104 (44.1%) | 316 (44.6%) |  |
| **Birth parent age (year)** | 31.2 (6.60) | 31.4 (6.99) | 31.3 (6.73) | 0.72 |
| **Gestational age (weeks)** | 38.4 (2.82) | 36.6 (4.64) | 37.8 (3.67) | <0.0001 |
| **Nulliparous** | 166 (35.2%) | 82 (34.7%) | 248 (35.0%) | 0.91 |
| **Infant sex (female)** | 239 (50.6%) | 118 (50.0%) | 357 (50.4%) | 0.87 |
| **Diagnosis** |  |  |  | NA |
| No genetic disease | 472 (100%) | 0 (0%) | 472 (66.7%) |  |
| Any genetic disease | 0 (0%) | 236 (100%) | 236 (33.3%) |  |
| Aneuploidy | 0 (0%) | 96 (40.7%) | 96 (13.6%) |  |
| Other chromosomal | 0 (0%) | 43 (18.2%) | 43 (6.1%) |  |
| Single-gene non-biochemical | 0 (0%) | 83 (35.2%) | 83 (11.7%) |  |
| Single-gene biochemical | 0 (0%) | 6 (2.5%) | 6 (0.8%) |  |
| Imprinting | 0 (0%) | 8 (3.4%) | 8 (1.1%) |  |

With the exposure of fetal genetic conditions defined, we performed an independent, double-coded chart review to assess factors associated with maternal health conditions during pregnancy and infant genetic disease, respectively. We conducted two separate assessments of the maternal chart (“double coding”) to ensure accuracy and consistency of the following abstracted data elements: presence or absence of preeclampsia, gestational hypertension, gestational diabetes, preexisting hypertension, preexisting diabetes, placental disorders, and obesity. When chart abstraction differed across assessors, these instances were reviewed, and a consensus determination was made following discussion with the study supervisor (JLC).

### Diagnosis Categories

The following maternal data elements were abstracted from the EHR: presence or absence of specific health conditions during pregnancy (see below), age at delivery, race, ethnicity, parity. Pregnancy complications were classified as follows: diabetes (preexisting type 1 or type 2 diabetes and gestational diabetes), hypertension or preeclampsia, and placental complications. A notation of abnormal glucose level without a diabetes diagnosis was coded as negative for gestational diabetes or pre-existing diabetes. If hypertension or diabetes was described as a disease ‘complicating pregnancy’, we considered this to be a preexisting version of that condition, rather than a gestational-onset condition. Premature rupture of membranes (PROM), pre-viability premature rupture of membranes (PPROM), Anti-Rho or other antibodies, and postpartum hemorrhage were not coded as health factors of concern. Conditions affecting the placenta were coded in the following categories: low-lying placenta/placenta previa; spectrum of placenta accreta, increta, percreta along with retained placenta; placental abruption; placental infarction/insufficiency; placentitis, and ‘other’ placental issues (which included circumvallate placenta and placenta chorangioma).

The following data elements were abstracted from the infant charts: singleton vs. twin or multiple gestation, sex, presence or absence of genetic diagnosis, and genetic diagnosis type, categorized as follows: aneuploidy, other chromosomal anomaly, single-gene biochemical, single-gene non-biochemical, and imprinting conditions.

### Statistical Analysis

All numerical data were evaluated using R Studio 2022.02.2 Build 485 Statistical Software using Package “readxl” R Statistical Software.^16,17^ To compare demographic attributes between exposed and unexposed dyads, a Pearson’s Chi-squared Test was used for categorical variables. Risk ratios (RR) were calculated by logistic regression using a log-link to determine the associations between pregnancy complications and fetal genetic conditions, adjusting for maternal age, parity, preexisting hypertension or diabetes, and obesity. RR were reported with their 95% confidence intervals.

## RESULTS

Of the 19,583 mothers who gave birth to children at Duke from 2013-2021, 18,809 had infants with no genetic consult or genetic testing, and 797 had infants receiving a genetic consult and/or genetic testing. Based on a review of the 797 charts in which infants underwent a genetic consult or testing, we identified 236 mothers of infants with a postnatally confirmed genetic diagnosis; these mother-infant dyads were considered the exposed group for the purposes of this study. Of the 18,809 mothers who had infants with no genetic consult or genetic testing, 472 were selected as matched dyads in the unexposed group (Figure 1, Table 1). Exposed dyads with confirmed infant genetic diagnoses were categorized into 5 groups: aneuploidy (N=96; 40.7%), other chromosomal anomalies (N=43; 18.2%), single-gene non-biochemical (N=83; 35.2%), single-gene biochemical (N=6; 2.5%), and imprinting condition (N=8; 3.4%) (Figure 2).

**Figure 1.**
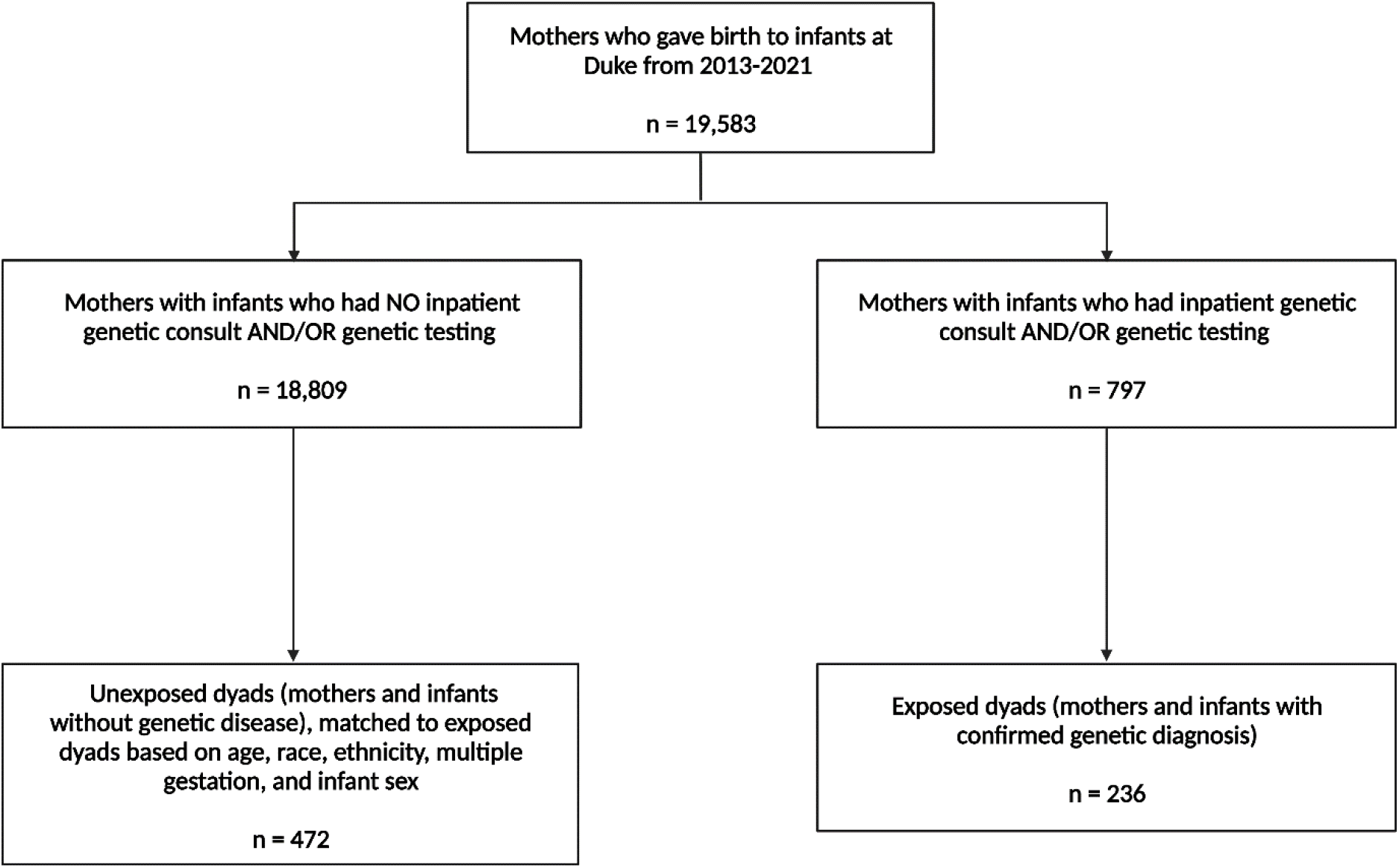
CONSORT diagram of retrospective cohort selection. Unexposed dyads included all mothers who gave birth to a child at Duke between 2013-2021 and did not receive a genetic consult or testing for their child. Exposed dyads included all mothers who gave birth to children at DUHS-affiliated hospitals between 2013-2021 and have a child with a confirmed genetic diagnosis.

**Figure 2.**
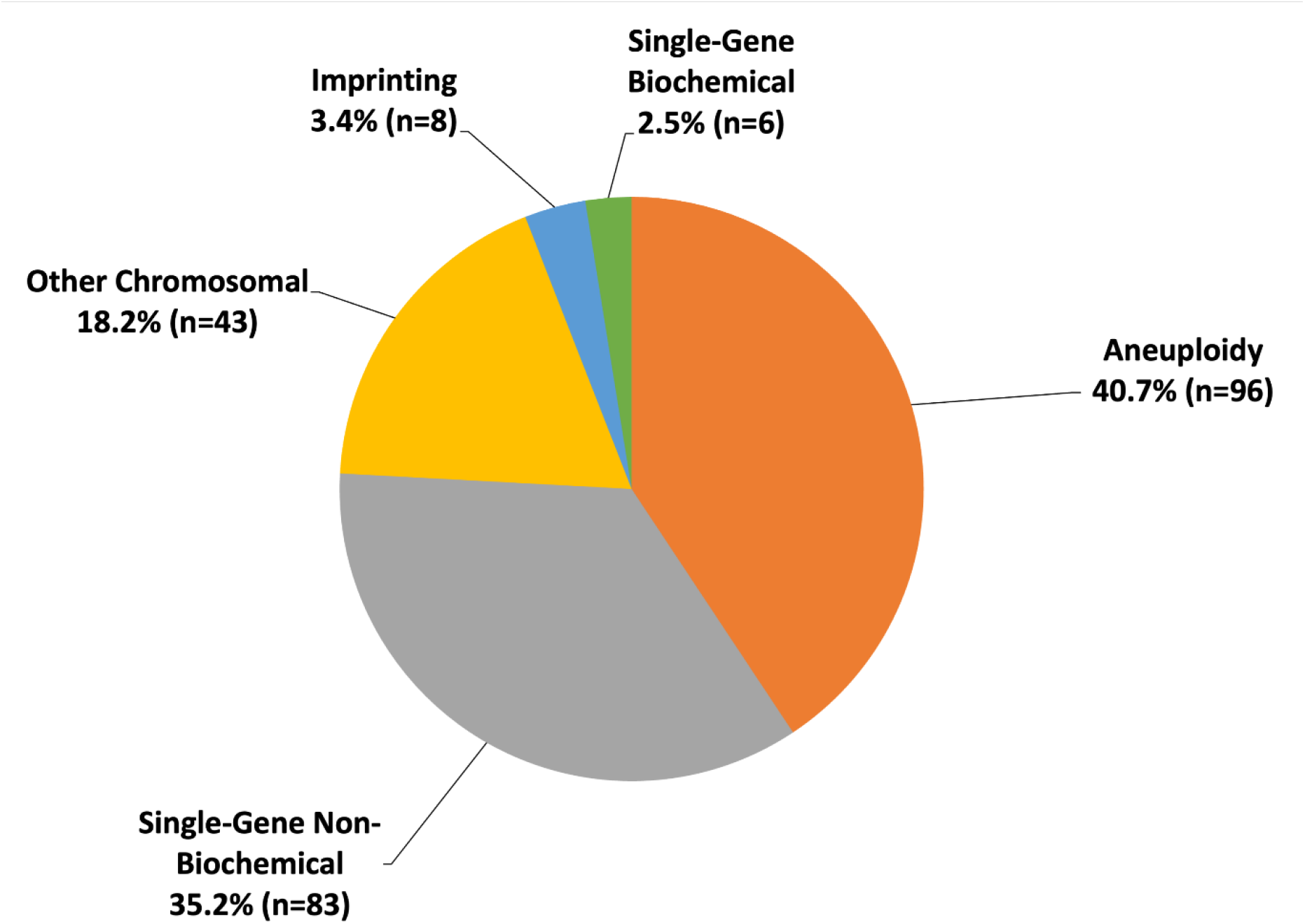
Distribution of genetic diagnoses across pregnancies affected by a fetal genetic disease. The exposed group included 236 mothers who had an infant with a confirmed genetic diagnosis. Of the diagnosed diseases, there were 96 cases classified as aneuploidy (40.3%), 43 cases classified as other chromosomal (18.9%), 83 cases classified as single-gene non-biochemical (35.2%), 6 cases classified as single-gene biochemical (2.5%), and 8 cases classified as imprinting (3.4%).

Pregnancy complications, including preeclampsia or gestational hypertension, gestational diabetes, and placental issues, were identified in 51.7% exposed dyads and 33.5% unexposed dyads. Multivariable logistic regression models evaluated the associated risk of pregnancy complications in the exposed group. Preeclampsia risk was only modestly elevated among the exposed group compared to the unexposed group overall (RR=1.32; 95% CI 0.88,1.98), even controlling for age, parity, and obesity (Table 2). Importantly, when genetic diagnoses were subcategorized, the exposed dyads in which the infant had an imprinting condition had a higher risk of preeclampsia compared to the unexposed dyads (RR=5.08; 95% CI 2.41,10.73) (Table 2). Risk of gestational diabetes was modestly elevated among the exposed group compared to the unexposed group overall (RR=1.43; 95% CI 0.97,2.10) (Table 2); however, in dyads in which the infant had aneuploidy, there was a more elevated risk of gestational diabetes compared to unexposed dyads (RR=1.65; 95% CI 1.05,2.61) (Table 2).

**Table 2.**
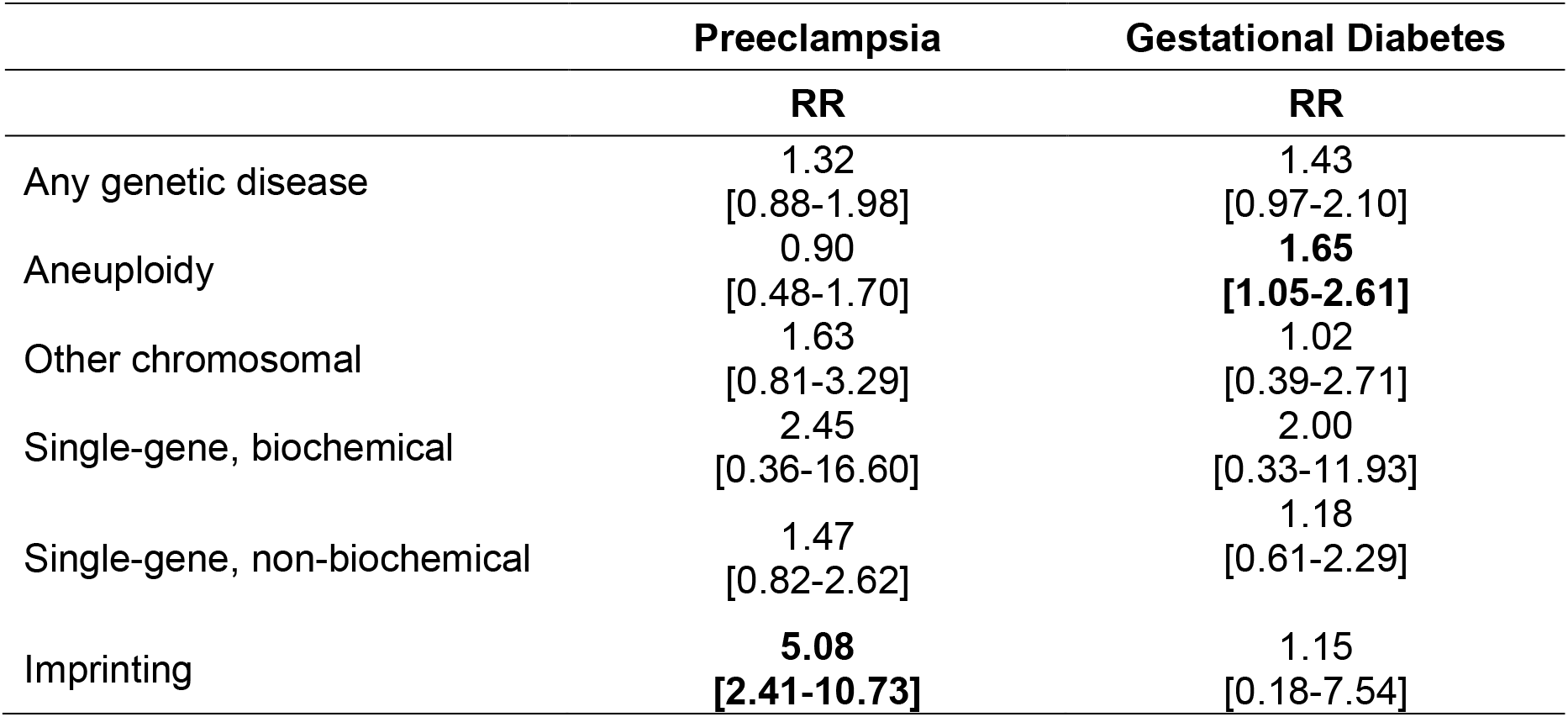
Preeclampsia and Gestational Diabetes risk in mothers of infants with genetic disease. Risk of preeclampsia and gestational diabetes were evaluated via multivariate logistic regression analyses. Risk Ratios were reported with 95% confidence intervals. The risk of preeclampsia was controlled for maternal age, parity, and obesity. The risk of gestational diabetes was controlled for maternal age and obesity. RR, risk ratio

Placental problems were identified in 27.1% exposed dyads and 10.2% unexposed dyads. Compared to the unexposed group, the exposed group had increased risk of any placental problem (RR 1.98; 95% CI 1.61,2.43), regardless of the type of genetic diagnosis in the infant (Table 3). We assessed associations between subcategories of placental problems and subcategories of infant genetic diagnosis. Of note, we removed subcategories with n<10 due to small sample size and the requirement to censor cells with fewer than 10 individuals. These omitted categories included imprinting conditions, single-gene biochemical conditions, low-lying placenta, and placentitis. Based on the stratified analysis of groups with n>10, the exposed group had an increased risk ratio of placental infarction for all reported genetic disease groups combined, including aneuploidy, other chromosomal anomalies, and single-gene non-biochemical genetic conditions (RR=2.25; 95% CI 1.72,2.93) (Table 3).

**Table 3.**
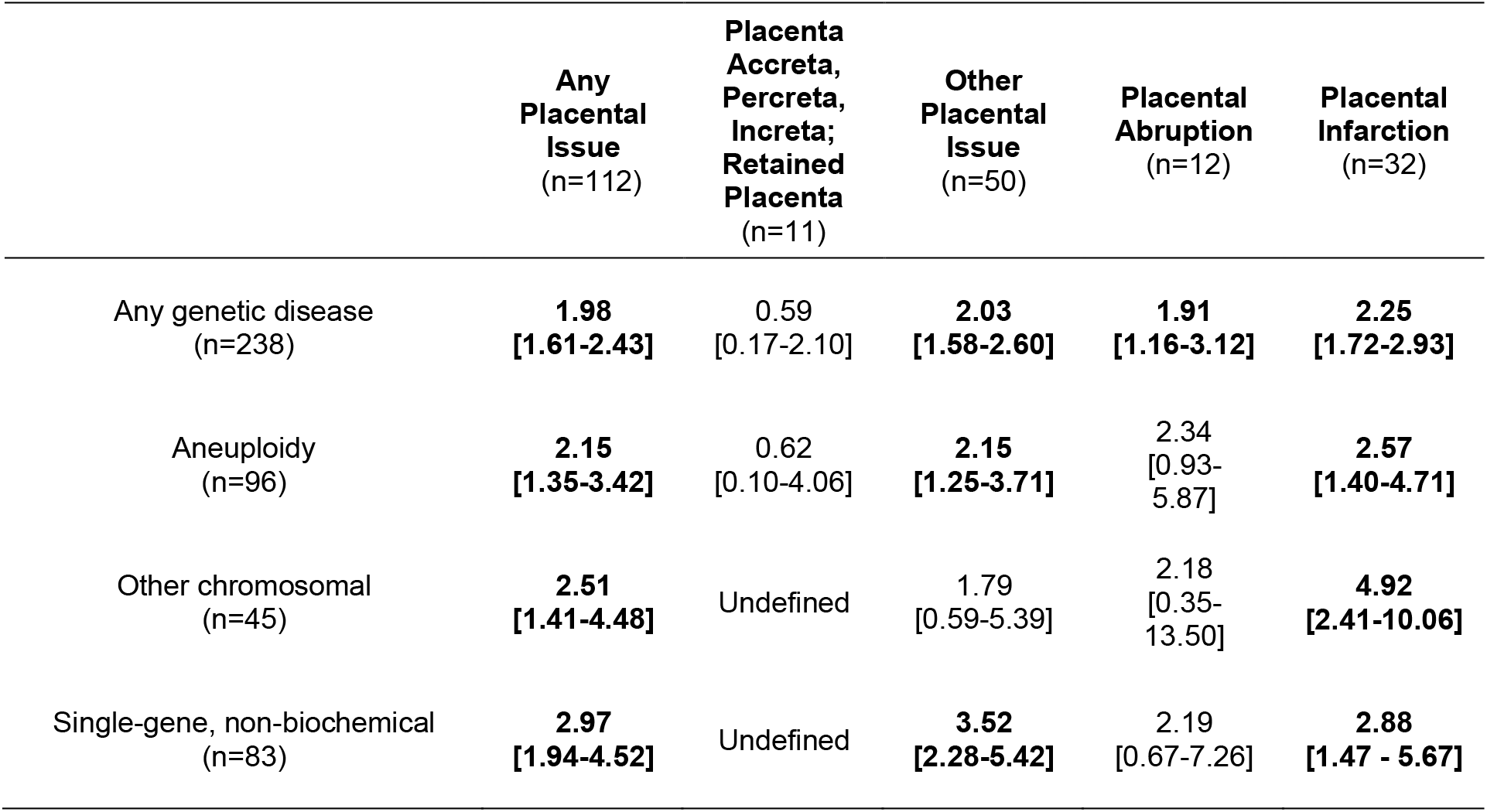
Placental disorders and associated risk in mothers of infants with genetic disease. Univariate logistic regression models were generated to answer the question of what, if any, associations there are with placental disorders and risk of genetic disease in offspring. Placental and genetic disease subcategories with n<10 were removed including imprinting, single-gene biochemical, low-lying placenta, and placentitis. Risk Ratios were reported with 95% confidence intervals.

|  | **Any**<br>**Placental**<br>**Issue**<br>**(n=112)** | **Placenta**<br>**Accreta,**<br>**Percreta,**<br>**Increta;**<br>**Retained**<br>**Placenta**<br>**(n=11)** | **Other**<br>**Placental**<br>**Issue**<br>**(n=50)** | **Placental**<br>**Abruptio**<br>**(n=12)** | **Placental**<br>**Infarction**<br>**(n=32)** |
| --- | --- | --- | --- | --- | --- |
| Any genetic disease<br>(n=238) | **1.98**<br>**[1.61-2.43]** | 0.59<br>[0.17-2.10] | **2.03**<br>**[1.58-2.60]** | **1.91**<br>**[1.16-3.12]** | **2.25**<br>**[1.72-2.93]** |
| Aneuploidy<br>(n=96) | **2.15**<br>**[1.35-3.42]** | 0.62<br>[0.10-4.06] | **2.15**<br>**[1.25-3.71]** | 2.34<br>[0.93-5.87] | **2.57**<br>**[1.40-4.71]** |
| Other chromosomal<br>(n=45) | **2.51**<br>**[1.41-4.48]** | Undefined | 1.79<br>[0.59-5.39] | 2.18<br>[0.35-13.50] | **4.92**<br>**[2.41-10.06]** |
| Single-gene, non-biochemical<br>(n=83) | **2.97**<br>**[1.94-4.52]** | Undefined | **3.52**<br>**[2.28-5.42]** | 2.19<br>[0.67-7.26] | **2.88**<br>**[1.47 - 5.67]** |

## COMMENTS

### Principal findings

We performed a retrospective chart review of maternal-infant dyads and used logistic regression to identify associations between fetal genetic diagnoses and pregnancy complications. Compared to pregnancies that were not affected by an infant with a genetic diagnosis, we found that risk of gestational diabetes was elevated 1.65-fold among pregnancies in which the infant was affected by aneuploidy and that preeclampsia risk was elevated 5-fold among pregnancies in which the infant was diagnosed with an imprinting condition. Furthermore, placental abnormalities were more common in pregnancies where the infant had any genetic disease diagnosis, including a notable 2-fold increase in placental infarction. More specifically, a significant increase in placental infarction was observed among pregnancies in which the infant was diagnosed with aneuploidy, other chromosomal anomalies, or single-gene non-biochemical genetic conditions. The associations between placental abnormalities and fetal genetic conditions were largely consistent in direction and magnitude across subgroups of genetic disease, although the increased risk ratio of placental infarction appeared largely attributable to the ‘other chromosomal anomaly’ subgroup, with a risk ratio of 4.92 (CI 1.47,10.06).

### Strengths of the study

Our study has several strengths and limitations. Strengths of the study include the relatively large size and diverse composition of our study sample, inclusion of unexposed dyads matched to exposed dyads by both demographics and parity, and the use of independent double coding to ensure accuracy and consistency of the chart review process.

### Limitations of the data

Limitations of the study include the retrospective, chart review-based data collection, the correlational nature of the study, and the use of data from a single site – specifically a large academic referral center. Our study did not match exposed and unexposed dyads by birth year; however, the year range in which infants were born in this study is relatively small and similar genetic testing was available throughout the study period. Our study also did not have data available to match exposed and unexposed dyads by social determinants of health, and choices may differ regarding continuing or terminating a pregnancy that is impacted by fetal genetic disease, based on social determinants of health. Because all genetic testing was done postnatally, prenatal genetic test results that led to elective termination of a pregnancy would not be captured as “exposed” dyads due to our study’s design.

### Interpretation of the data

Gestational diabetes is estimated to affect up to 10% of pregnancies worldwide, making it one of the most common pregnancy complications.^18^ Importantly, gestational diabetes is associated with multiple adverse birth outcomes, including fetal malformations, preeclampsia, macrosomia, and preterm birth.^19^ We identified an association between fetal aneuploidy and gestational diabetes, primarily Trisomy 13, 18, and 21 (OMIM # 190685). A prior study by Moore and colleagues also observed increased gestational diabetes in aneuploid pregnancies and specifically noted a higher prevalence among those with a sex chromosome difference.^1^ We did not identify a similar association; however, the number of infants with sex chromosome differences was relatively few within our cohort.

Prior literature has reported conflicting associations between specific fetal genetic diagnoses and pregnancy complications, particularly between fetal aneuploidy and preeclampsia or gestational hypertension.^2–8^ We did not observe fetal aneuploidy (consisting mainly of Trisomy 13, 18, and 21 in our cohort) to be strongly associated with elevated preeclampsia risk; nor did we did identify a general association between fetal genetic disease and risk of preeclampsia or gestational hypertension in our study. Interestingly, we identified an increased risk of gestational diabetes among pregnancies in which the fetus was diagnosed with aneuploidy, and an increased risk of preeclampsia among pregnancies in which the fetus was diagnosed with an imprinting disorder. Although pregnancies impacted by an imprinting disorder comprised a small percentage of pregnancies in our study, this finding warrants future investigation. Prior studies have designated specific biochemical genetic conditions or monogenic non-biochemical conditions as being associated with preeclampsia,^9–14^ but either the small numbers of these biochemical conditions in our study, or the fact that we grouped them into a category of disease rather than investigating a single disease in isolation may have impacted our ability to replicate these prior findings.

An important finding from our work was that all fetal genetic diseases in aggregate were associated with placental problems. We observed a significantly greater number of placental problems in pregnancies in which the fetus had any genetic disease diagnosis compared to pregnancies in which the fetus did not have a genetic disease diagnosis. Specifically, risk of placental infarction, ‘other placental issues,’ and all placental disorders combined was elevated among pregnancies affected by any fetal genetic diagnosis compared to those unaffected by fetal genetic disease. There was an association between placental issues and all subcategories of genetic disease, even when controlling for preeclampsia or preexisting hypertension. These findings are in line with a murine model study by Perez-Garcia and colleagues that characterized placental defects among embryonic lethal and sub-viable mouse knockout lines.^20^ In particular, Perez-Garcia and colleagues observed specific associations between placental conditions and variants in genes required for normal development of the heart, brain, and vascular system, suggesting potential molecular links between placenta-related health conditions and fetal genetic conditions. These findings support the need for future studies evaluating associations between specific fetal genetic alterations and placental defects to clarify the developmental mechanisms that are impaired (*e*.*g*., neovascularization, immunologic rejection, embryological patterning).

### Conclusions

The findings presented here may have utility in guiding medical management of mothers and infants at risk of a genetic disorder due to maternal heterozygote status or following a prenatal genetic testing result. For example, if a pregnancy is determined to be impacted by fetal aneuploidy, the medical care of the mother could benefit from augmented surveillance for gestational diabetes. Similarly, if preeclampsia or placental infarction is diagnosed, this may alert providers to have a lower threshold to evaluate the fetus or infant for genetic disease. It will be necessary to investigate whether directional causality exists among the associations identified in this and similar studies, including specific evaluation of the underlying biology linking fetal genetic conditions to pregnancy complications. Such studies will support the development of novel methods to diagnose, manage, and potentially prevent adverse gestational and birth outcomes associated with fetal genetic disease. Further, these findings will help support future work to develop algorithms for the prospective identification of infants potentially affected by a genetic condition. Such algorithms will be needed to guide the timing and deployment of expanded genetic testing methods, allowing early identification and treatment of infants affected by genetic disease, even prenatally. Early identification will be particularly important as new prenatal treatment modalities and expanded screening methods become available.

## Data Availability

The data that support the findings of this study are available upon request by emailing the corresponding author, subject to any restrictions regarding the need to restrict access to protected health information.

## AUTHOR CONTRIBUTIONS

RAG, BA, LE, KW, JH, and JLC conceived and designed the study. RAG, BA, JH, and JLC curated the data. RAG, BA, KW, JH, and JLC conducted the data analysis. RAG, BA, LE, JH, and JLC drafted the initial version of the manuscript. All authors provided important insight during the data analysis and critically revised the manuscript.

## ACKNOWLEDGEMENTS

This research was supported by the YT and Alice Chen Pediatric Genetics and Genomics Research Center at Duke University. The funding sources had no role in the study design, data collection and analysis, decision to publish, or preparation of the manuscript. The opinions in this article belong to the authors. Figure 1 was created using BioRender.com.

## CONFLICT OF INTEREST STATEMENT

The authors have no competing financial interests.

## FUNDING STATEMENT

This research was supported by the YT and Alice Chen Pediatric Genetics and Genomics Research Center at Duke University. The funding sources had no role in the study design, data collection and analysis, decision to publish, or preparation of the manuscript. The opinions in this article belong to the authors.

## ETHICS DECLARATION

This study was reviewed and approved as an exempt, retrospective research protocol by the DUHS Institutional Review Board, including a waiver of consent for retrospective chart review (protocol Pro00106469). All study activities were conducted in accordance with the Declaration of Helsinki.

## Notes

### Competing Interest Statement

The authors have declared no competing interest.

### Author Declarations

The Duke University Health System (DUHS) Institutional Review Board deemed this retrospective study exempt from review (protocol number Pro00106469).

